# Both vascular risk factors and common genetic variants influence penetrance of variants causing monogenic stroke

**DOI:** 10.1101/2022.05.24.22275509

**Authors:** Bernard PH Cho, Eric L Harshfield, Maha Al-Thani, Daniel J Tozer, Steven Bell, Hugh S Markus

**Author notes:** Correspondence to: Prof. Hugh Stephen Markus, University of Cambridge, Department of Clinical Neurosciences, Neurology Unit, R3, Box 83, Cambridge Biomedical Campus, Cambridge CB2 0QQ. contributed equally (Steven Bell, Hugh S Markus).

## Abstract

Monogenic forms of stroke have been thought to be rare with high penetrance. However, recent studies have reported typical monogenic stroke pathogenic variants are much commoner than expected in the general population. Whether such variants are associated with disease, and why the phenotype of these variants varies so widely remain unclear. In 454,787 individuals in UK Biobank, we identified typical pathogenic variants in *NOTCH3, HTRA1* and *COL4A1/2* genes in 1 in 467, 1 in 832 and 1 in 1353 subjects, respectively. Variants in all three genes were associated with stroke risk, and *NOTCH3* and *HTRA1* with dementia risk. Cardiovascular risk (assessed by Framingham cardiovascular risk score), polygenic risk (assessed by a polygenic stroke risk score), and variant location within each gene, were all associated with penetrance of *NOTCH3* and *HTRA1* variants. Our results suggest intensive cardiovascular risk factor modification may reduce stroke and dementia risk in individuals with such variants.

## INTRODUCTION

A number of monogenic or single gene disorders are associated with increased risk of stroke and vascular dementia. These predominantly cause the small vessel subtype of ischaemic stroke (lacunar stroke), associated with damage to the small perforating arterials supplying the white matter and deep grey matter nuclei. The most common of these monogenic disorders is Cerebral Autosomal Dominant Arteriopathy with Subcortical Infarcts and Leukoencephalopathy (CADASIL) caused by variants in *NOTCH3*, with the second most frequent being CADASIL2 caused by autosomal dominant variants in *HTRA1*. Variants in *COL4A1/2* can also cause small vessel stroke, and intracerebral haemorrhage.

Such monogenic forms of stroke have been thought to be very rare with an estimated prevalence of CADASIL of 4 per 100,000 the United Kingdom.^1^ However, recent studies of large exome and whole genome sequencing databases have reported that pathogenic variants are much more frequent in the general population,^2^ with typical *NOTCH3* variants being present in 1 in 452 individuals,^3^ and pathogenic *HTRA1* variants reported in 1 in 275.^4^

These novel and unexpected findings raises two important questions. Firstly, are these apparently ‘asymptomatic’ variants associated with stroke and dementia? Secondly, why do some individuals who have these variants present with severe early onset stroke and dementia, while others remain apparently symptom free?

Initial studies from UK Biobank, in which exome sequencing data is available in combination with phenotypic information including long-term outcomes, reported that these *NOTCH3* variants were associated with stroke and MRI features of small vessel disease, particularly white matter hyperintensities.^3^ Another study in 17,800 individuals reported similar associations for *HTRA1*.^4^ These studies used data derived from a subset of 200,000 UK Biobank participants, but exome sequencing data is now available on the majority of the cohort, allowing significantly increased power to detect associations with disease.

Until recently, it was thought that CADASIL was a disease with high penetrance and that most individuals with typical variants would suffer early onset stroke. More recently, the clinical phenotype in CADASIL cases with typical *NOTCH3* variants has been shown to be widely variable, with some individuals suffering early onset stroke while other remain stroke free at least until the eighth decade.^5^ A number of factors have been suggested to modulate the phenotype including variant location (with variants in proximal EGF repeats 1-6 being associated with more severe disease),^2^ the presence of cardiovascular risk factors,^6^ and modifying genes.^7^ The UK Biobank resource also includes data on vascular risk factors and imputed genome-wide genotyping, allowing cardiovascular risk factor and polygenic risk scores to be calculated to investigate the importance of such modifying factors.

To explore this area further, we analysed the latest UK Biobank cohort with exome sequencing data. We identified pathogenic variants in the three most common monogenic forms of small vessel disease caused by variants in *NOTCH3, HTRA1*, and *COL4A1/2*. We determined the frequency of such variants and their associations with both prevalent and incident stroke and dementia. We also determined associations with MRI markers of cerebral small vessel disease, namely white matter hyperintensities (WMH) and more diffuse white matter damage detected on diffusion tensor imaging (DTI).

## METHODS

### Study population

UK Biobank is a prospective study of over 500,000 healthy volunteers aged 40–69 years recruited across the UK between 2006 and 2010.^8^ About 9 million individuals were invited to join, of whom 5.5% participated in the baseline assessment.^9^ Phenotypic data were collected through questionnaires and physical examinations. A subset of 100,000 individuals underwent MRI scans, of which approximately 47,000 brain MRIs were available at the time of this analysis. These participants were selected on the basis of travelling distance from the imaging centre and not clinical information. All MRIs were performed on one of the three identical Skyra 3.0T scanners (Siemens Medical Solutions, Germany). Identical acquisition parameters and quality control were used for all scans.^10^ In October 2021, whole-exome sequences of 454,787 UK Biobank participants were released and all were included here. About 38,000 of the 47,000 with brain MRI had exome sequencing data available.

### Ascertainment of pathogenic variants

The variants located in the *NOTCH3* (chromosome 19:15,159,038-15,200,995, reference genome assembly GRCh38), *HTRA1* (chromosome 10:122,458,551-122,514,907), *COL4A1* (chromosome 13:110,148,963-110,307,157) and *COL4A2* (chromosome 13:110,305,812-110,513,209) genes were extracted from the exome data in PLINK format. The extracted variants were annotated using Ensembl Variant Effect Predictor.^11^ These variants were then filtered according to an *a priori* set of pathogenicity criteria for each of the genes (as specified below).

For *NOTCH3*, we identified all CADASIL variants that result in the gain or loss of a cysteine residue in one of the 34 EGFR domains of the NOTCH3 protein (amino acid position 40-1373, Uniprot accession number: Q9UM47).

Pathogenic variants in *HTRA1 and COL4A1/2* are not stereotyped in the way *NOTCH3* variants are. Therefore, we performed a systematic review to identify those that had been reported in patients with familial cerebral small vessel disease. We searched PubMed for the following terms (CARASIL OR HTRA1 mutation*) AND (COL4A1 mutation* OR COL4A2 mutation* OR COL4A1/2 mutation*) and selected publications in English up to and including 5 February 2022. These variants were classified using the American College of Medical Genetics and Genomics (ACMG) criteria for classifying pathogenic variants,^12^ with the support of the Association for Clinical Genomic Science (ACGS) Best Practice Guidelines for Variant Classification in Rare Disease 2020.^13^ Notably, statistical evaluation of co-segregation of variant with disease (PP4) was performed using the thresholds suggested by a previous study;^14^ and criterion PM2 was upgraded to strong for any glycine-changing variants that affect the triple helix region of COL4A1/2.^13^ Binary (yes/no) results for all the ACMG criteria were combined to classify variants as “pathogenic”, “likely pathogenic” or “uncertain”. Finally, only the variants that were classified as “pathogenic” or “likely pathogenic” were included in the subsequent analyses.

### Phenotypic data fields

History of vascular risk factors, measured blood pressure, blood pressure medication and parental history of stroke were recorded. History of diseases, including migraine, migraine with aura, any stroke, ischaemic stroke, intracerebral haemorrhage, vascular dementia, all-cause dementia and epilepsy, were determined from self-report, hospital and death records (code list in **supplementary table 1**). Cases of stroke, ischaemic stroke, intracerebral haemorrhage and vascular dementia diagnosed after recruitment to UK Biobank were identified as incident cases.

### Brain imaging analysis

In the 38,332 participants with exome sequences and MRI available, measures were compared between variant subjects and controls. Among the sequences acquired were T1-weighted images, fluid attenuated inversion recovery (FLAIR) images and DTI images, full details of the acquisition were previously reported,^15^ brief details are given in **supplementary table 2**. We used derived MRI measures for brain volume and WMH volume, generated by an image-processing pipeline developed and run on behalf of UK Biobank.^10^ Brain volume was estimated from the T1-weighted images by SIENAX,^16^ and normalised for head size. WMHs were quantified on FLAIR images through the brain intensity abnormality classification algorithm,^17^ and normalised for brain volume. The degree of white matter ultrastructural damage assessed on the DTI data was calculated using automated software to derive the peak width skeletonised mean diffusivity (PSMD),^18^ a measure of the width of the MD histogram from the centre of the white matter tracts, which provides a summary measure from the DTI data. Additionally, DTI data was used to derive structural brain networks via tractography analysis,^19^ from which global and local structural efficiency measures^20^ were derived in-house.^21^ Following tractography, the automated anatomic labelling atlas^22^ was used to generate a connectivity matrix using the ninety non-cerebellar regions of the atlas. More details were previously reported.^23^ Global efficiency concerns integration over the whole network and is estimated by averaging efficiency for all node pairs (i.e. how many steps it takes to go from any given node to any other). It reflects the ability to rapidly combine information from different brain regions. The average local efficiency measures clustering/segregation and specialization within a network - that is, the capability to perform specialised processing within densely interconnected brain regions - and is calculated from the efficiency of the connections between first-degree neighbours of each node (i.e. the global efficiency of a sub-graph defined by a given node and those nodes connected to it). Global network efficiency has been shown to be sensitive to damage in cerebral small vessel disease, correlate with cognitive impairment,^24^ and predict future dementia risk.^25^

### Calculation of Framingham cardiovascular and polygenic risk scores

To compare the risk of ischaemic stroke conferred by *NOTCH3* and *HTRA1* variants with that conferred by cardiovascular risk factor burden and that conferred by common genetic stroke variants, we calculated both the Framingham cardiovascular risk score (FRS)^26^ and an ischaemic stroke polygenic risk score (PRS) using more than 3 million common genetic variants,^27^ respectively.

When calculating the HRs for ischaemic stroke associated with 1-SD increase in FRS and PRS as well as presence of *NOTCH3* and *HTRA1* variants, Cox proportional hazards regression models were limited to participants without a history of stroke, coronary heart disease, peripheral vascular disease or congestive heart failure at recruitment. By dividing the HR associated with the variant status by that associated with 1-SD higher FRS or PRS (on the log-scale, assuming a linear association), we estimated the increment of the FRS and PRS in SD that was predicted to be equivalent to the risk conferred by *NOTCH3* and *HTRA1* variants.^28^ We performed statistical tests for interaction between the FRS or PRS and variant status of each gene; for this we divided our sample cohort into two groups based on their FRS or PRS: low (bottom 50%) and high (top 50%) risk.^29^ Multiplicative and additive interaction was assessed by ANOVA and the synergy index, respectively.^30^

### Statistical analysis

The effects of *NOTCH3, HTRA1* and *COL4A1/2* variants on phenotypes were assessed by linear regression for continuous outcomes and logistic regression for binary outcomes. For the former, effect sizes were standardised to enable more informative comparisons. A heterozygous carrier of *NOTCH3* and *HTRA1* variants were excluded from our analyses. Firth’s correction was applied to all logistic regression models to account for rare event bias.^31^ All regression models were adjusted for age, sex, ethnicity, exome sequencing batch and the first 10 principal components of genetic ancestry. WMH volumes were natural log-transformed for analyses. Kaplan-Meier analyses and Cox proportional hazards regression (using time since recruitment to UK Biobank as the underlying timescale) were performed to compare the cumulative probability of incident disease in different variant groups and in different tiers of PRS and FRS. All statistical analyses were performed using R v.4.0.3 with two-sided p-values and p <0.05 considered statistically significant.

## RESULTS

### Systematic review and classification of *HTRA1* and *COL4A1/2* variants

Our initial search for *HTRA1* variants yielded 391 publications (**figure 1**). After reviewing the abstracts, 70 articles were selected for review of the full article, of which 21 were excluded after full review. Reasons for exclusion include lack of variant information and duplicate cases. The remaining 49 articles were assessed for patient’s phenotype and their variants. 77 unique *HTRA1* variants were found in 135 patients. There were 30 homozygous cases (63% in Asia), 103 heterozygous cases (58% in Europe) and 2 compound heterozygous cases (one in China and one in Europe).

**Figure 1.**
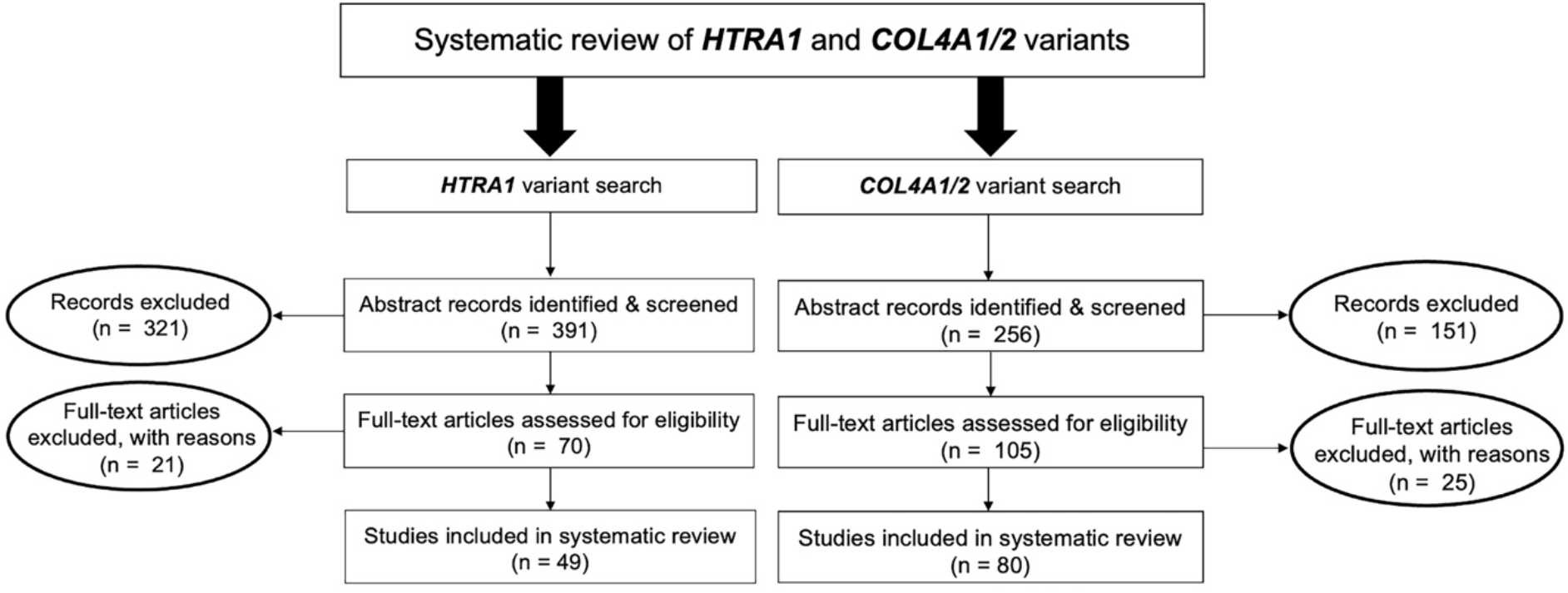
Flow chart of literature search and selection of studies for *HTRA1* and *COL4A1/2* variants.

For *COL4A1/2*, 295 articles were initially identified (**figure 1**), 105 of which were selected. After full review, 25 of these studies were excluded with reasons such as lack of variant information, variant in patients with unconfirmed cerebral small vessel disease (without brain MRI or CT scans) and duplicate cases. The remaining 80 articles were assessed for patient’s phenotype and their variants. 164 unique *COL4A1* variants were identified in 275 patients, and 34 *COL4A2* variants in 51 patients. There was a pair of homozygous siblings for each gene.

After ACMG classification, we found 63 pathogenic (n=28) or likely pathogenic (n=35) variants in *HTRA1*, 131 (100 pathogenic and 31 likely pathogenic) in *COL4A1*, and 21 (4 pathogenic and 17 likely pathogenic) in *COL4A2*. 44.4% of the *HTRA1* variants were missense and affected the protease domain. Although an *HTRA1* variant (p.Gln151Lys) was classified as likely pathogenic using the ACMG criteria, we excluded it from our analyses based on recent functional evidence which indicated it does not affect protease activity.^4^ 81% and 89% of the *COL4A1* and *COL4A2* variants, respectively, were glycine-changing and affected the triple helix region. Overall, the ACMG criteria excluded about 20-40% of the clinically reported variants with uncertain pathogenic significance.

### Prevalence and distribution of *NOTCH3, HTRA1* and *COL4A1/2* pathogenic variants

Of the 454,787 participants with exome sequencing data, 973 (2.1 per 1000) carried a *NOTCH3* pathogenic variant. All the carriers were heterozygotes, giving a population frequency of 1 in 467. In total, 99 unique *NOTCH3* pathogenic variants were identified, of which 54 were previously reported on dbSNP,^32^ and 49 were observed in only one individual (**supplementary figure 1 and supplementary table 3**). The identified variants were spread across 22 different exons between exons 2 and 24 of the *NOTCH3* gene. They affected 33 EGFR domains, involving all EGFR domains except 21. Variants were predominantly located towards the distal end of the NOTCH3 protein; only 28 participants (3%) had a *NOTCH3* variant located in EGFR domains 1-6. Two of the most common *NOTCH3* pathogenic variants in the cohort, p.Arg1231Cys and p.Cys1222Gly, together accounted for 48% of all NOTCH3 variant carriers (n=467).

For *HTRA1*, 546 participants (1.2 per 1000) carried a pathogenic variant. All the carriers were heterozygotes, giving a population frequency of 1 in 832. In total, 18 unique *HTRA1* pathogenic variants were identified, of which 4 were carried by only one individual (**supplementary figure 2 and supplementary table 4**). Two-thirds of the identified variants affected the protease domain, and these variants were carried by 92% of all the *HTRA1* variant carriers. Notably, one of the *HTRA1* variants, p.Arg227Trp, was found in 379 participants, accounting for 69% of the *HTRA1* variant carriers.

For *COL4A1/2*, 336 participants carried a pathogenic variant (196 with *COL4A1* and 140 with *COL4A2*). All carriers were heterozygotes, giving a population frequency of 1 in 1353. In total, 11 unique pathogenic variants (8 in *COL4A1* and 3 in *COL4A2*) were identified (**supplementary figures 3 & 4 and supplementary table 5**). All the identified variants affected the triple helix region, of which 9 were glycine-changing, 1 was missense (p.Pro352Leu) and 1 was nonsense (p.Arg1063Ter). Of note, p.Gly332Arg was the most common variant in *COL4A1*/2 and found in 174 individuals.

### Association between *NOTCH3, HTRA1, COL4A1/2* variants and prevalent stroke, vascular dementia, and other clinical features

The presence of a *NOTCH3* variant was associated with at least twofold increase in the odds of any stroke (odds ratio [OR]: 2.16, 95% confidence interval [CI]: 1.67 to 2.74, p=3.2×10^−8^), ischaemic stroke (OR: 2.65, 95% CI: 1.96 to 3.50, p=5.9×10^−9^) and intracerebral haemorrhage (OR: 2.42, 95% CI: 1.23 to 4.22, p=0.01). All-cause dementia (OR: 2.26, 95% CI: 1.52 to 3.23, p=0.0001) and vascular dementia (OR: 5.42, 95% CI: 3.11 to 8.74, p=2.7×10^−7^) were also more prevalent in variant carriers. Population attributable fractions showed that 0.21% of strokes and 0.77% vascular dementia in UK Biobank were attributed to the *NOTCH3* variants. There was also an increase in the odds of epilepsy (OR:1.72, 95% CI: 1.12 to 2.51, p=0.01). Presence of a variant was associated with an increased family history of stroke (OR:1.50, 95% CI: 1.31 to 1.71, p=6.0×10^−9^). No significant associations were found with migraine (OR: 1.21, 95% CI: 0.89 to 1.61, p=0.22) or migraine with aura (OR: 1.57, 95% CI: 0.18 to 5.62, p=0.61) (**figure 2**).

**Figure 2.**
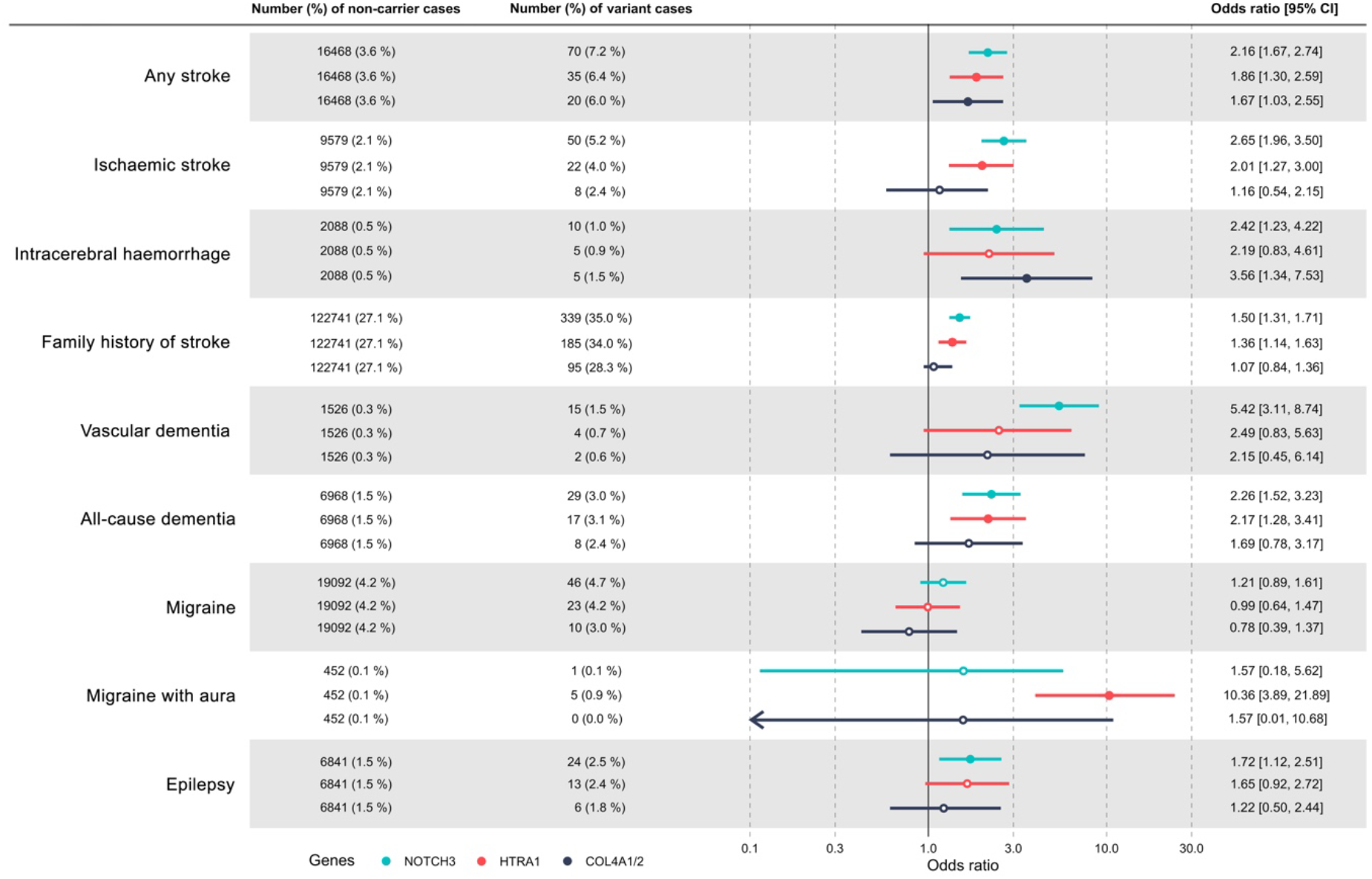
Association of *NOTCH3, HTRA1* and *COL4A1/2* variants with lifetime history of different cerebral small vessel disease related diagnoses. Firth’s correction was applied to all the regression models which were adjusted for age, sex, ethnicity, exome sequencing batch and the first 10 genetic ancestry principal components. N=454,276.

The presence of an *HTRA1* variant was associated with an increased likelihood of migraine with aura (OR: 10.36, 95% CI: 3.89 to 21.89, p=7.6×10^−5^), any stroke (OR: 1.86, 95% CI: 1.30 to 2.59, p=0.001) and ischaemic stroke (OR: 2.01, 95% CI: 1.27 to 3.00, p=0.004). All-cause dementia (OR: 2.17, 95% CI: 1.28 to 3.41, p=0.005) was more prevalent in variant carriers. 0.09% of strokes and 0.14% vascular dementia in UK Biobank were attributed to the *HTRA1* variants. Carriers were also more likely to have a family history of stroke (OR:1.36, 95% CI: 1.14 to 1.63, p=0.001). No significant association was found with vascular dementia (OR:2.49, 95% CI: 0.83 to 5.63, p=0.10) or any other clinical outcomes (**figure 2**).

The presence of a *COL4A1/2* variant was associated with an increased risk of any stroke (OR: 1.67, 95% CI: 1.03 to 2.55, p=0.04), which was accounted for by a marked increase in intracerebral haemorrhage risk (OR: 3.56, 95% CI: 1.34 to 7.53, p=0.01), while there was no difference in ischaemic stroke risk (OR: 1.16, 95% CI: 0.54 to 2.15, p=0.69). 0.04% of strokes in UK Biobank were attributed to the *COL4A1/2* variants (**figure 2**).

Variant carrier prevalence and associations with clinical outcomes were consistent after limiting our analyses to unrelated individuals (data not shown).

### Association between variants and MRI features of SVD

MRI data were available for 92 *NOTCH3*, 44 *HTRA1* and 28 *COL4A1/2* variant carriers. *NOTCH3* variants were associated with increased WMH volume (standardised difference: 0.48, 95% CI: 0.32 to 0.64, p=5.5×10^−9^) and increased DTI assessed white matter ultrastructural damage on PSMD (standardised difference: 0.64, 95% CI: 0.46 to 0.83, p=8.5×10^−12^), as well as a decrease in local (standardised difference: -0.25, 95% CI: -0.45 to -0.05, p=0.01) and global (standardised difference: -0.27, 95% CI: -0.47 to -0.07, p=0.01) structural efficiency (**table 1**). They were associated with an increase in brain volume (standardised difference: 0.24, 95% CI: 0.07 to 0.40, p=0.01).

*HTRA1* variants were associated with an increase in WMH volume (standardised difference: 0.55, 95% CI: 0.31 to 0.78, p=5.6×10^−6^) and DTI-PSMD (standardised difference: 0.68, 95% CI: 0.42 to 0.95, p=5.2×10^−7^), and a decrease in local (standardised difference: -0.48, 95% CI: -0.76 to -0.20, p=0.001) and global (standardised difference: -0.50, 95% CI: -0.77 to -0.22, p=0.0004) structural efficiency (**table 1**). They were not associated with brain volume changes.

**Table 1.** Standardised effects of *NOTCH3, HTRA1, COL4A1/2* variants on different MRI markers.

| Genes | Markers | Non-variant carriers | Variant carriers | Standardised effect [LCI, UCI] | p |
| --- | --- | --- | --- | --- | --- |
| ***NOTCH3*** | Brain volume | 39,598 | 92 | 0.24 [0.07, 0.40] | 0.01 |
|  | WMH volume | 38,295 | 89 | 0.48 [0.32, 0.64] | <0.01 |
|  | PSMD | 37,274 | 87 | 0.64 [0.46, 0.83] | <0.01 |
|  | Local efficiency of structural brain network | 34,486 | 84 | -0.25 [-0.44, -0.05] | 0.01 |
|  | Global efficiency of structural brain network | 34,486 | 84 | -0.27 [-0.47, -0.07] | 0.01 |
| ***HTRA1*** | Brain volume | 39,405 | 44 | 0.21 [-0.03, 0.45] | 0.08 |
|  | WMH volume | 38,110 | 42 | 0.55 [0.31, 0.78] | <0.01 |
|  | PSMD | 37,094 | 42 | 0.68 [0.41, 0.95] | <0.01 |
|  | Local efficiency of structural brain network | 34,313 | 42 | -0.48 [-0.76, -0.20] | <0.01 |
|  | Global efficiency of structural brain network | 34,313 | 42 | -0.50 [-0.77, -0.22] | <0.01 |
| ***COL4A1/2*** | Brain volume | 39,598 | 28 | 0.14 [-0.16, 0.44] | 0.35 |
|  | WMH volume | 38,295 | 23 | -0.01 [-0.33, 0.30] | 0.93 |
|  | PSMD | 37,274 | 24 | 0.06 [-0.29, 0.41] | 0.73 |
|  | Local efficiency of structural brain network | 34,486 | 24 | 0.10 [-0.27, 0.47] | 0.59 |
|  | Global efficiency of structural brain network | 34,486 | 24 | 0.11 [-0.25, 0.48] | 0.55 |

Measurements of brain volume, white matter hyperintensities (WMH), peak width skeletonised mean diffusivity (PSMD) as well as global and local efficiency of structural brain network were analysed with the presence of the variants through linear regression with adjustment for age at scan, ethnicity, sex, sequencing batch and genetic principal components. LCI, lower confidence interval; UCI, upper confidence interval. Brain and WMH volumes were natural-log transformed.

*COL4A1/2* variants were not associated with any of the MRI markers (**table 1**).

### Association between variant carriers and incident stroke and vascular dementia

During follow-up for cardiovascular and dementia end-points for a median duration of 12.6 years (25^th^-75^th^ percentiles, 11.8-13.2), 6.6% (64 out of 972) of *NOTCH3* variant carriers and 3.4% (15,314 out of 452,864) of non-carriers (those without any of the *NOTCH3, HTRA1* and *COL4A1/2* variants) had an incident stroke. *NOTCH3* variant status was a predictor of incident stroke (hazard ratio (HR) for *NOTCH3* carriers versus non-carriers: 1.94, 95% CI: 1.51 to 2.48, p=1.3×10^−7^) (**figure 3A**). Incident vascular dementia occurred in 1.3% (13 out of 972) of *NOTCH3* variant carriers and 0.3% (1269 out of 452,864) of non-carriers. Cox regression showed *NOTCH3* variant status was associated with incident vascular dementia after controlling for confounding factors (HR: 4.74, 95% CI: 2.74 to 8.19, p=2.5×10^−8^) (**figure 3B**).

**Figure 3.**
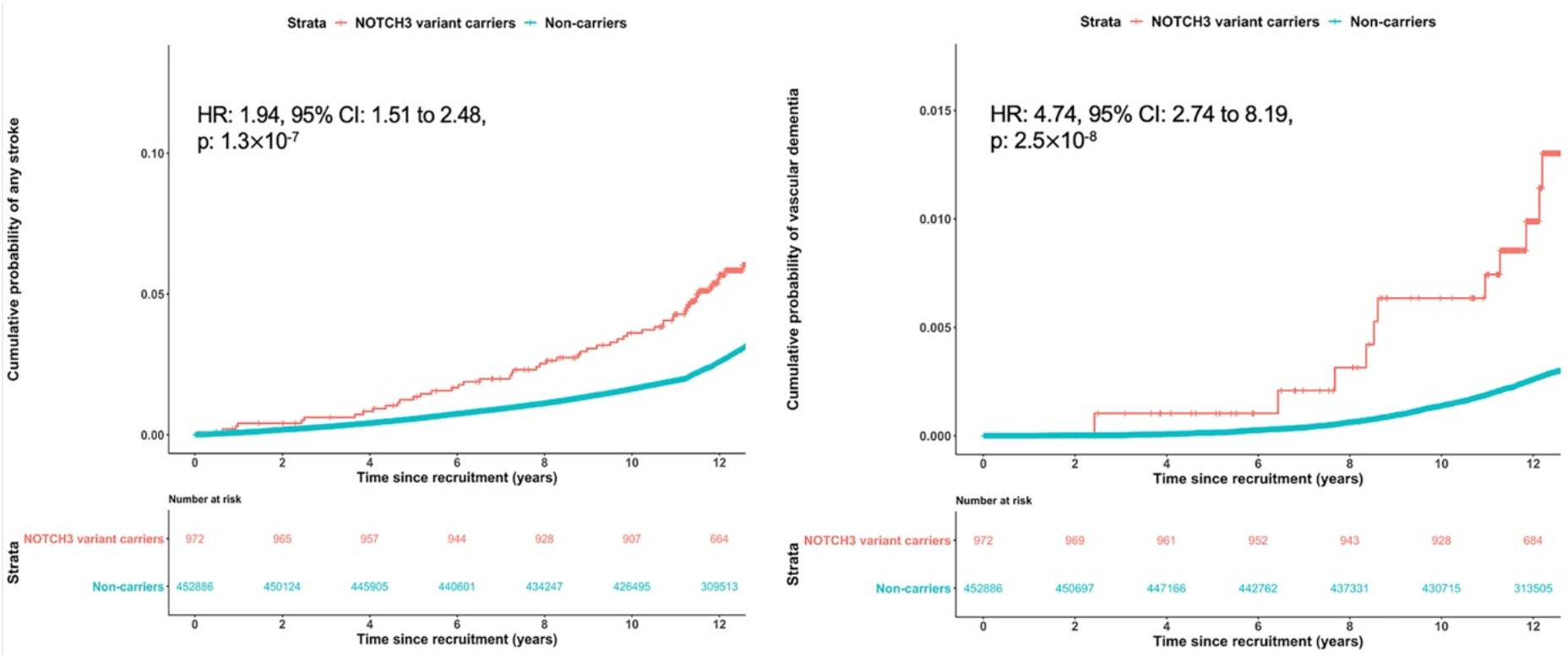
Cumulative probability of (A, left) any stroke and (B, right) vascular dementia in individuals with and without a *NOTCH3* pathogenic variant. Hazard ratio (HR) was calculated through Cox regression with Firth’s correction and adjustment for sex, ethnicity, exome sequencing batch and the first 10 genetic ancestry principal components.

5.7% (31 out of 545) of *HTRA1* variant carriers had incident stroke, and 0.6% (3 out of 545) had incident vascular dementia. *HTRA1* variant status was associated with incident stroke (HR for *HTRA1* carriers versus non-carriers: 1.70, 95% CI: 1.19 to 2.42, p=0.003) but not with vascular dementia (HR: 2.07, 95% CI: 0.67 to 6.43, p=0.21). For *COL4A1/2*, 5.4% (18 out of 336) of the variant carriers had incident stroke, and 0.6% (2 out of 336) had incident vascular dementia. *COL4A1/2* variant status was not predictive of incident stroke (HR for *COL4A1/2* carriers versus non-carriers: 1.38, 95% CI: 0.87 to 2.18, p=0.18) or vascular dementia (HR: 1.80, 95% CI: 0.45 to 7.23, p=0.40).

### Effect of modulating factors on phenotype of the whole cohort

#### Location of variants

##### Position of NOTCH3 variants

22 of the *NOTCH3* variant carriers had a variant in EGFRs 1-6. Compared to EGFR 7-34 variant carriers, carriers of the EGFRs 1-6 variant had an increased risk of migraine (OR:6.61, 95% CI: 2.14 to 17.89, p=0.002), any stroke (OR:14.01, 95% CI: 5.34 to 36.56, p=6.6×10^−7^), ischaemic stroke (OR:13.78, 95% CI: 4.92 to 36.73, p=5.7×10^−6^), vascular dementia (OR:82.68, 95% CI: 22.49 to 358.03, p=4.9×10^−10^) and all-cause dementia (OR:46.19, 95% CI: 14.24 to 162.62, p=3.0×10^−9^) but not of epilepsy (OR:2.17, 95% CI: 0.23 to 9.46, p=0.42) (**table 2**).

**Table 2.** Odds ratios and 95% confidence intervals for the association of variant position in *NOTCH3, HTRA1, COL4A1/2* with lifetime diagnosis of different cerebral small vessel disease associated diseases.

| Medical Records | NOTCH3 EGFRs<br>1-6 vs 7-34<br>N<sub>1</sub>=22 vs N<sub>2</sub>=950 | p.Arg227Trp vs<br>other *HTRA1*<br>variants<br>N<sub>1</sub>=378 vs N<sub>2</sub>=167 | COL4A1:p.Gly332Arg<br>vs other *COL4A1/2*<br>N<sub>1</sub>=174 vs N<sub>2</sub>=162 |
| --- | --- | --- | --- |
| Any stroke | 14.01 [5.34, 36.56] | 0.39 [0.20, 0.79] | 1.72 [0.61, 5.15] |
| Ischaemic stroke | 13.78 [4.92, 36.73] | 0.28 [0.11, 0.65] | 1.10 [0.23, 5.65] |
| ICH | 10.39 [1.78, 43.32] | 0.59 [0.12, 3.38] | 5.72 [0.73, 102.93] |
| Family history of stroke | 8.55 [3.23, 27.99] | 1.21 [0.81, 1.81] | 0.75 [0.45, 1.26] |
| Vascular dementia | 82.68 [22.49, 358.03] | 0.21 [0.03, 1.20] | 25.46 [0.73, 3144929.33] |
| All-cause dementia | 46.19 [14.24, 162.62] | 0.43 [0.16, 1.20] | 2.74 [0.58, 15.87] |
| Migraine | 6.61 [2.14, 17.89] | 0.92 [0.39, 2.35] | 2.20 [0.59, 11.70] |
| Migraine with aura | 4.31 [0.03, 96.82] | 0.17 [0.02, 0.90] | No cases |
| Epilepsy | 2.17 [0.23, 9.46] | 0.77 [0.27, 2.52] | 0.52 [0.06, 3.12] |

Firth's correction was applied to all the regression models which were adjusted for age, sex, ethnicity, exome sequencing batch and the first 10 principal components of genetic ancestry.

##### Position of HTRA1 variants

379 of the *HTRA1* variant carriers had a variant at one location (p.Arg227Trp). Compared to the rest of the *HTRA1* carriers, carriers of the p.Arg227Trp variant had a lower risk of any stroke (OR:0.39, 95% CI: 0.20 to 0.79, p=0.01), ischaemic stroke (OR:0.28, 95% CI: 0.11 to 0.65, p=0.003). They had similar risk of any migraine (OR:0.92, 95% CI: 0.39 to 2.35, p=0.85), but a lower risk of migraine with aura (OR:0.17, 95% CI: 0.02 to 0.90, p=0.04) (**table 2**).

##### Position of COL4A1/2 variants

172 of the *COL4A1/2* variant carriers had a variant at p.Gly332Arg. Compared to the rest of the *COL4A1/2* carriers, carriers of the p.Gly332Arg variant did not differ in their risk of any of the clinical outcomes we assessed (**table 2**).

#### Cardiovascular risk profile

We next calculated the effect of vascular risk factors on ischaemic stroke risk in individuals with and without variants. For this analysis, we confined ourselves to *NOTCH3* and *HTRA1* as significant associations with ischaemic stroke and dementia had only been found for these variants.

Cardiovascular risk factor burden, as assessed by the FRS, increased the risk of ischaemic stroke associated with both *NOTCH3* and *HTRA1* variants (HR for a 1-SD increase in FRS: 1.40, 95% CI: 1.36 to 1.45, p<0.001; **figure 4**). No evidence for a multiplicative interaction was observed. However, there was an additive interaction between FRS and *NOTCH3* (p-value for the synergy index = <0.01) and *HTRA1* (synergy index p-value = <0.01) carrier status. We calculated *NOTCH3* variants conferred the same risk as a 2.84 SD increase in FRS and *HTRA1* variants a 1.73 SD increase. With our most conservative estimates, these placed carriers of *NOTCH3* and *HTRA1* variants as having a cardiovascular risk equivalent to the upper 4.5% and 45.8% of the population, respectively.

**Figure 4.**
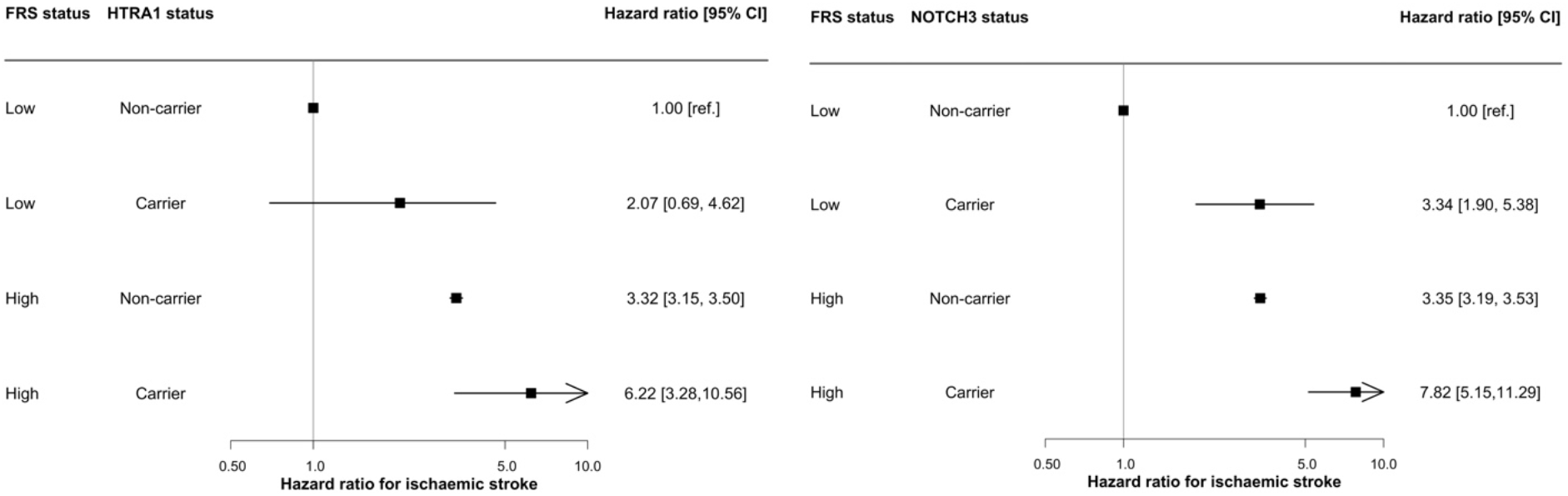
Hazard ratios and associated 95% confidence intervals for ischaemic stroke stratified by high and low Framingham cardiovascular risk score and (A, left) *NOTCH3* or (B, right) *HTRA1* status. (A) P-value for a multiplicative interaction with the categorical low and high FRS groupings = 0.07; p-value for the additive interaction with categorical FRS, assessed using the synergy index <0.01. (B) P-value for a multiplicative interaction with the categorical FRS groupings = 0.85; p-value for the additive interaction with categorical FRS, assessed using the synergy index <0.01.

#### Polygenic risk

Again focussing on *NOTCH3* and *HTRA1* as for the FRS, common genetic variants, as assessed by the polygenic risk score, increased the risk of stroke (HR for 1-SD increase in PRS: 1.27, 95% CI: 1.24 to 1.31, p<0.001; **figure 5**). This was seen to a similar extent both in those with and without variants, and unlike the FRS, there was no additive interaction between PRS and *NOTCH3* (synergy index p-value = 0.18) and *HTRA1* (synergy index p-value = 0.28) carrier status. We calculated *NOTCH3* variants conferred the same risk as a 3.94 SD increase in PRS and *HTRA1* variants a 2.40 SD increase. With our most conservative estimates, these placed carriers of *NOTCH3* and *HTRA1* variants as having a polygenic risk equivalent to the upper 1.0% and 44.2% of the population, respectively.

**Figure 5.**
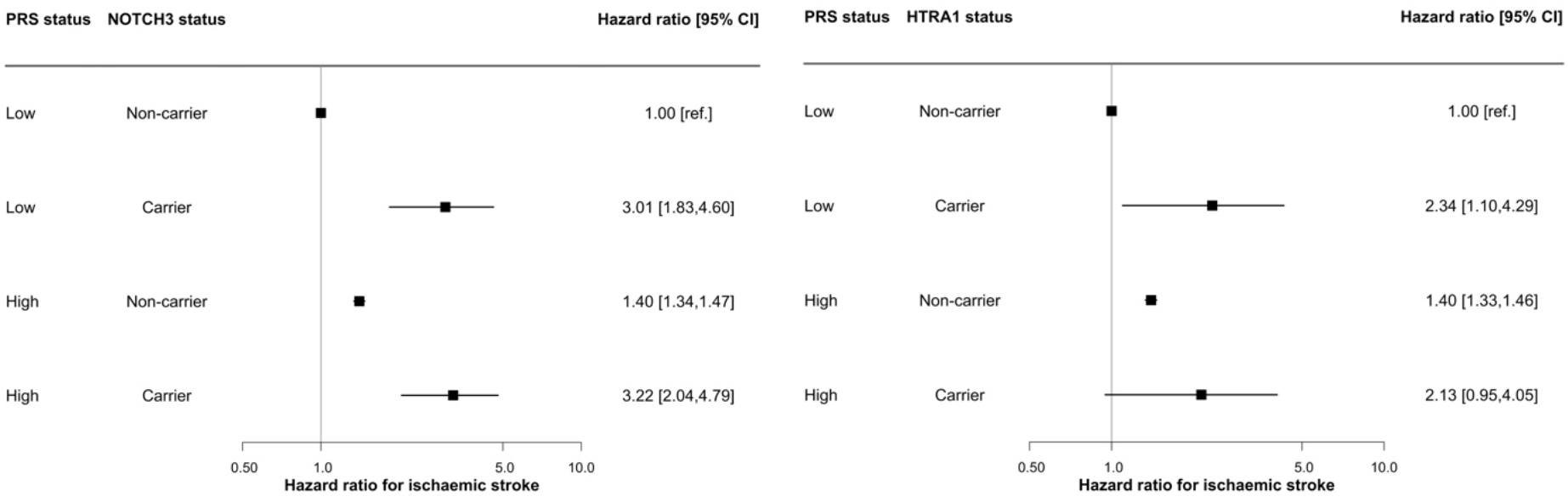
Forest plot showing the hazard ratios and associated 95% confidence intervals for ischaemic stroke stratified by high and low polygenic risk score and (A, left) *NOTCH3* or (B, right) *HTRA1* status. (A) P-value for a multiplicative interaction with the categorical low and high PRS groupings = 0.97; p-value for the additive interaction with categorical PRS, assessed using the synergy index = 0.18. (B) P-value for a multiplicative interaction with the categorical PRS groupings = 0.29; p-value for the additive interaction with categorical PRS, assessed using the synergy index = 0.28.

## DISCUSSION

In this study of over 450,000 individuals, we demonstrated the frequency of pathogenic variants in three genes associated with monogenic small vessel disease, specifically *NOTCH3, HTRA1*, and *COL4A1/2*, and that such variants are much more common than the clinical monogenic disease caused by variants in these genes. This is consistent with recent reports both from a subset of UK Biobank participants and other sequencing databases,^3,4,33,34^ and demonstrated that such variants occur in between 2 and 3 per 1000 of the UK Biobank population. Our results extend previous work on a subset of UK Biobank by providing definitive data on the association of such variants with disease. Specifically, we demonstrated that both *NOTCH3* and *HTRA1* variants were associated with increased risk of all stroke, ischaemic stroke, and vascular dementia. Risk of intracerebral haemorrhage was also elevated in those carrying pathogenic *NOTCH3* variants. In contrast, although *COL4A1/2* variants were associated with increased risk of any stroke, this was accounted for by a marked increase in intracerebral haemorrhage risk and no significant difference in ischaemic stroke risk was found.

The frequency of such variants raises the question as to why many individuals with these variants remain asymptomatic. This contrasts with patients presenting with familial forms of SVD, such as CADASIL and CADASIL2, in which most family members tend to have symptoms at some time in their life. This has led to the clinical impression that CADASIL was highly penetrant. Our results suggest that site of the variant within the gene, conventional cardiovascular risk factors, and polygenic risk, play a role in determining penetrance. For both *NOTCH3* and *HTRA1* variants, in whom there were sufficient ischaemic stroke cases to perform the analysis, cardiovascular risk factors, as assessed by the FRS, increased risk of ischaemic stroke in individuals carrying pathogenic genetic variants, and there was a statistical interaction between variant status and FRS. This is consistent with previous cross-sectional data from much smaller cohorts of symptomatic CADASIL patients in which both smoking and hypertension were associated with increased stroke risk.^35,36^ Genetic propensity to common ischaemic stroke, as assessed by an ischaemic stroke PRS, increased ischaemic stroke risk in individuals with *NOTCH3* and *HTRA1* variants, but the extent of the increase was similar that seen in those without variants, and there was no interaction between PRS and variant status. This is consistent with data from other diseases demonstrating that common genetic variants affect the penetrance of monogenic conditions,^37^ and also with data from symptomatic CADASIL cases in which white matter lesion volume was demonstrated to have significant heritability after controlling for variant site.^7^

We also demonstrated that variant location had an impact on disease severity. Previous studies have demonstrated that proximal *NOTCH3* variants in EGFR 1-6 are associated with more severe disease,^2,3^ and our data confirmed this with a markedly increased risk of both stroke and dementia in individuals with proximal variants. We extended this finding to show similar effects in *HTRA1* where one variant (p.Arg227Trp) was found to have less severe disease than the lower risk of stroke.

The availability of approximately 40,000 MRI scans allowed us to not only examine associations with clinical disease, but also determine associations with MRI markers of small vessel disease. This analysis showed that both *NOTCH3* and *HTRA1* variants were associated with increased WMH volume, reduced white matter integrity as assessed by PSMD, and disruption of structural brain networks as assessed by local and global efficiency measures. Structural network measures have been shown to mediate the effect of other SVD pathologies, such as WMH and lacunar infarcts, on cognition.^24^ Surprisingly, *NOTCH3* variants were associated with an increase in brain volume and a similar trend was found for *HTRA1*. The explanation for this is unclear as patients with severe symptomatic CADASIL have atrophy and reduced brain volume. It is possible that in earlier stages of the disease pathological changes are associated with inflammatory or oedematous changes that could increase brain volume although this area requires further research.

The strengths of our study are that we were able to study the largest cohort to date (450,000 individuals) and that we extended previous work to analyse associations with all three of the most common monogenic forms of SVD. Furthermore, the prospective data collected within UK Biobank allowed us to compare risk of incident stroke conferred by variants to that from a risk prediction score used in clinical practice. Our study also has limitations. The study sample was large but is not necessarily representative of the wider UK population as there may be a selection bias in who agreed to participate. However, one would not expect this to affect the inferences in this study. Secondly, in UK Biobank we only had endpoints of ischemic and hemorrhagic stroke but not specifically with lacunar stroke and we were therefore unable to examine associations of variants directly with lacunar stroke.

In summary, we have demonstrated in 450,000 individuals that relatively common variants in genes causing monogenic SVD increase the risk of both stroke and dementia. This increase in risk is primarily via ischaemic stroke for *NOTCH3* and *HTRA1*, but via an increase in intracerebral haemorrhage for *COL4A1/2*. Our results show that factors contributing to the wide variation in penetrance seen include variant location within the gene itself, conventional vascular risk factors and polygenic risk. An implication of our results is that intensive vascular risk factor control is likely to improve disease prognosis in individuals with these genetic variants. This raises the possibility that identifying individuals early in life prior to onset of symptoms and the occurrence of covert MRI changes of SVD might help reduce the risk of stroke and dementia.

## Data Availability

Data from UK Biobank (https://www.ukbiobank.ac.uk/) are available to bona fide researchers on application.

## FUNDING

This research was funded by the British Heart Foundation via the Cambridge British Heart Foundation Centre of Research Excellence (PhD studentship awarded to BPHC and fellowship awarded to ELH) (RE/18/1/34212) and a British Heart Foundation programme grant (RG/4/32218). Infrastructural support was provided by the Cambridge University Hospitals NIHR Biomedical Research Centre (BRC-1215-20014). HSM is supported by a NIHR Senior Investigator Award. The views expressed are those of the authors and not necessarily those of the NIHR or the Department of Health and Social Care.

## AUTHOR CONTRIBUTION

B.P.H.C., S.B. and H.S.M. conceived and designed the study. B.P.H.C., M.A. and S.B. performed and reviewed the systematic review of *HTRA1* and *COL4A1/2* variants. B.P.H.C., E.L.H. and S.B. extracted and analysed the genetic and phenotypic data. B.P.H.C. and D.J.T. downloaded and analysed the neuroimaging data. B.P.H.C, S.B. and H.S.M. interpreted the data. B.P.H.C. and H.S.M. wrote the first draft of the manuscript. All authors contributed to writing the final version of the manuscript.

## COMPETING INTERESTS

The authors report no competing interests.

## DATA AVAILABILITY

Data are available in a public, open access repository. Data from UK Biobank (https://www.ukbiobank.ac.uk/) are available to bona fide researchers on application. This study was performed under UK Biobank application number 36509.

## Notes

### Competing Interest Statement

The authors have declared no competing interest.

### Author Declarations

UK Biobank received ethical approval from the NHS National Research Ethics Service North West (21/NW/0157).

