## supplementary table 2 and supplementary figures 1-4 for "Both vascular risk factors and common genetic variants influence penetrance of variants causing monogenic stroke"

**SUPPLEMENTARY INFORMATION**
**(5 tables and 4 figures)**

**Supplementary table 1 [Separate file, eTable_1.xlsx].** The code list used for the retrieval of disease records.

**Supplementary table 2.** An overview of the brain MRI scanner and its sequence parameters.

| **Scanner** | **Siemens Skyra 3T** |
|---|---|
| **T1-weighted structural imaging** | **Straight sagittal orientation** (i.e. with the field-of-view aligned to the scanner axes)<br>**TR** (repetition time)=2000 ms; **TE** (echo time)=2.01 ms<br>**Resolution**:1x1x1 mm; **Field-of-view**: 208x256x256 matrix<br>**Duration**: 5 minutes<br>**Others:** 3D magnetization-prepared rapid acquisition with gradient echo (MPRAGE), in-plane acceleration iPAT=2, prescan-normalise |
| **T2-weighted FLAIR structural imaging** | **Straight sagittal orientation**<br>**TR**=5000 ms; **TE**=395 ms<br>**Resolution**: 1.05x1x1 mm; **Field-of-view**: 192x256x256 matrix;<br>**Duration**: 6 minutes<br>**Others**: 3D sampling perfection with application-optimized contrasts by using flip angle evolution (SPACE), in-plane acceleration integrated Parallel Acquisition Techniques (iPAT)=2, partial Fourier = 7/8, fat saturation, elliptical k-space scanning, prescan-normalise |
| **Diffusion imaging** | **TR**=3600 ms; **TE**=92 ms<br>**Resolution**: 2x2x2 mm; **Field-of-view**: 104x104x72 matrix<br>**Duration**: 7 minutes (including 36 seconds phase-encoding reversed data), 5x b=0 (+3x b=0 blip-reversed), 50x b=1000 s/mm$^2$, 50x b=2000 s/mm$^2$<br>**Gradient timings**: duration=21.4 ms, spacing = 45.5 ms; Spoiler b-value = 3.3 s/mm$^2$<br>**Number of gradient directions:** For the two diffusion-weighted shells, 50 distinct diffusion-encoding directions were acquired (and all 100 directions are distinct). The diffusion preparation is a standard ("monopolar") Stejskal-Tanner pulse sequence.<br>**Others:** Spin-echo echo planar imaging (SE-EPI) with x3 multislice acceleration, no iPAT, fat saturation |

**Supplementary table 3 [Separate file, eTable_3.xlsx].** Information about the ninety-nine distinct cysteine-altering *NOTCH3* variants.
**Supplementary table 4 [Separate file, eTable_4.xlsx].** Information about the eighteen *HTRA1* pathogenic variants. Details of the ACMG classification are also included.
**Supplementary table 5 [Separate file, eTable_5.xlsx].** Information about the eleven *COL4A1/2* pathogenic variants. Details of the ACMG classification are also included.

**Supplementary figure 1. Lolliplot showing the distribution of distinct pathogenic variants in UK Biobank across the reverse strand of the *NOTCH3* gene.** Blue circles represent variants affecting EGFRs 7-34; red circles represent variants affecting EGFRs 1–6. EGFR, epidermal growth factor-like repeat.

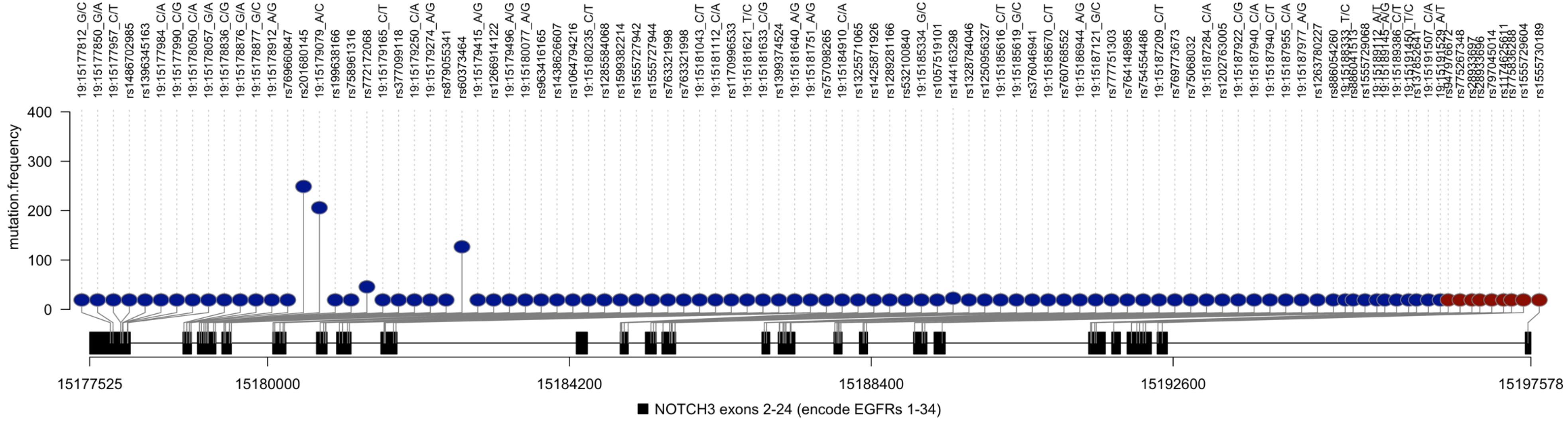


**Supplementary figure 2. Lolliplot showing the distribution of distinct pathogenic variants in UK Biobank across the forward strand of the *HTRA1* gene.** Red represents variants affecting the protease domain; blue represents other variants; circle represents missense variants; inverted triangle represents a nonsense variant.

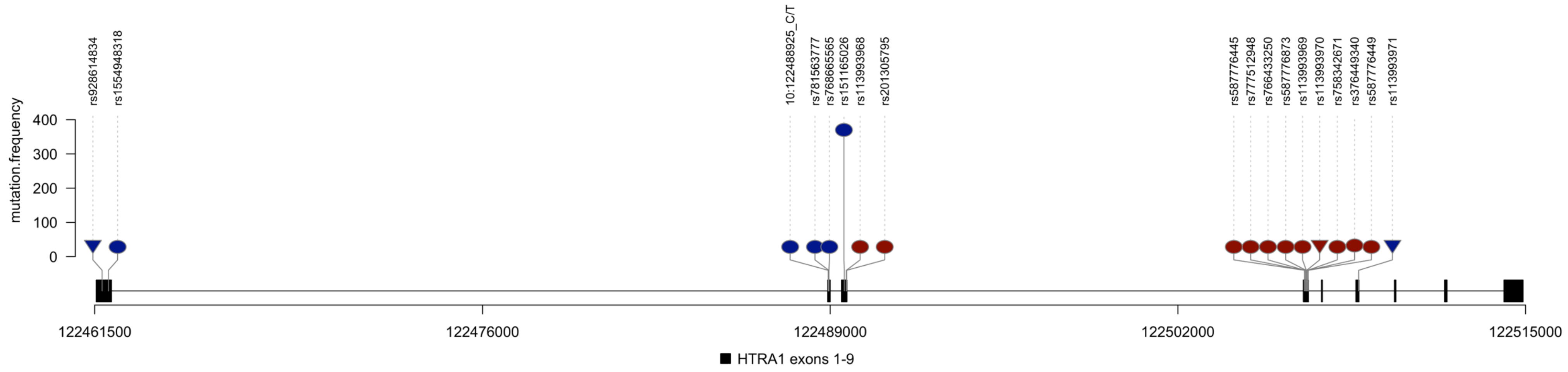

**Supplementary figure 3. Lolliplot showing the distribution of distinct pathogenic variants in UK Biobank across the reverse strand of the *COL4A1* gene.** Red represents glycine-changing variants affecting the triple helix region; blue represents other variants; circle represents missense variants; inverted triangle represents a nonsense variant.

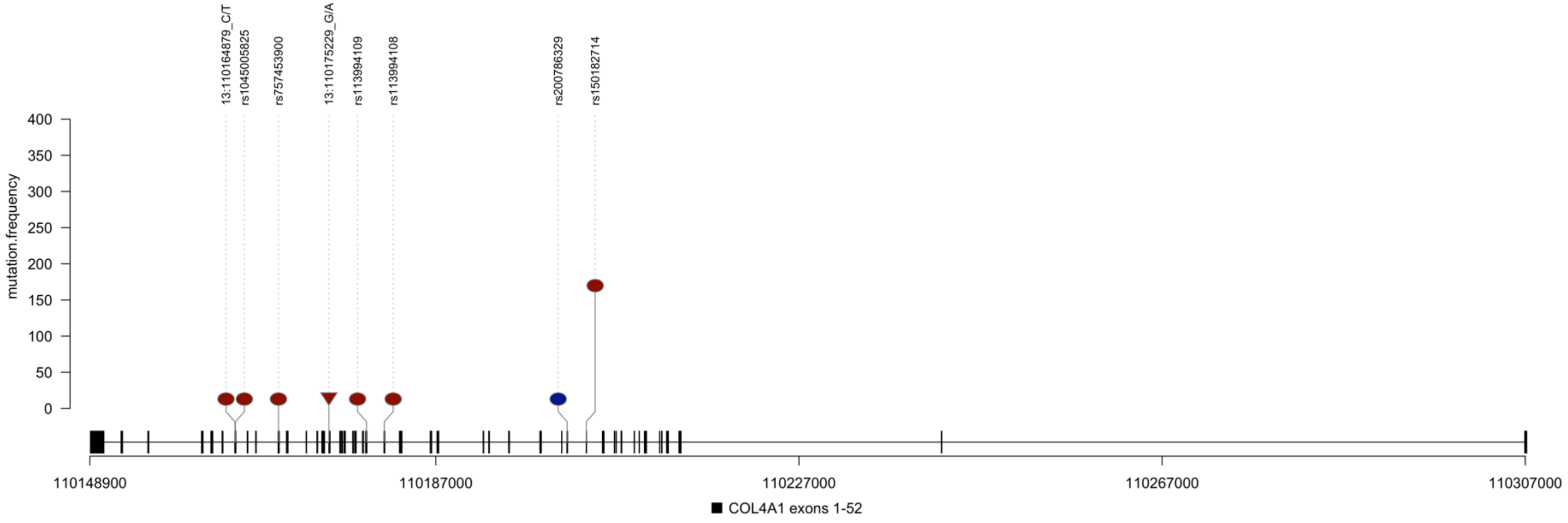


**Supplementary figure 4. Lolliplot showing the distribution of distinct pathogenic variants in UK Biobank across the forward strand of the *COL4A2* gene.** Red circle represents glycine-changing variants affecting the triple helix region.

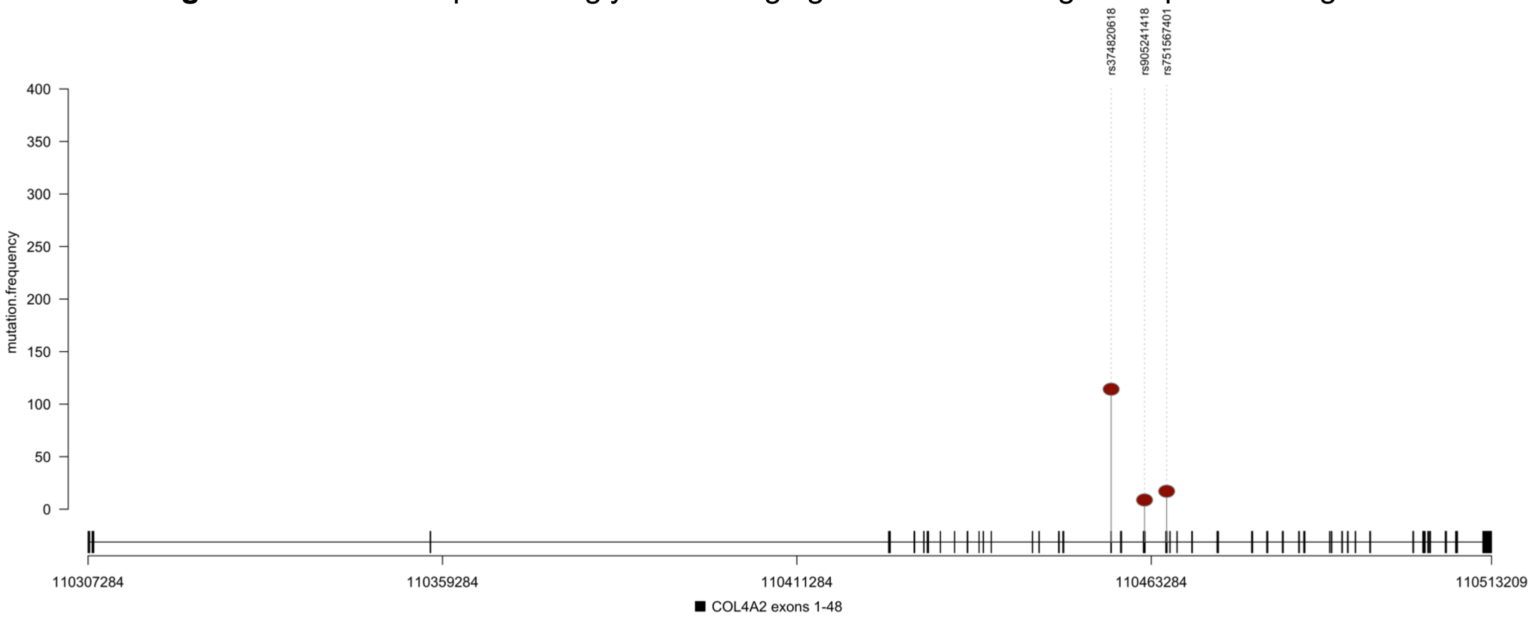
